# Radiofrequency Monitoring of Intracranial Hemorrhage in the Neurological Intensive Care Unit: A Pilot Trial of the SENSE Device

**DOI:** 10.1101/2020.05.25.20109108

**Authors:** Joseph J. Korfhagen, George J. Shaw, Matthew L. Flaherty, Opeolu Adeoye, William A. Knight

## Abstract

Intracerebral hemorrhage (ICH) is a devastating form of stroke, with substantial mortality and morbidity. Hemorrhage expansion (HE) occurs in ~40% of patients and portends worse neurologic outcome and mortality. Currently, ICH patients are admitted to the intensive care unit (ICU) and monitored for HE with neurologic exam and neuroimaging. By the time a change is detected, it may be too late to mitigate HE. There is a clinical need for a non-invasive bedside monitor of ICH.

The SENSE consists of a 9-antenna array mounted around the head with driving electronics. A 913 MHz signal is transmitted by a given antenna, crosses the intracranial region, and received by the remaining 8 antennae. A complete measurement consists of one cycle with each antenna serving as the transmitting antenna. It was hypothesized that HE of ≥ 3 ml would be detected by the device.

Ten ICH subjects admitted within 24 hours of stroke onset were enrolled. All patients received a diagnostic head CT (baseline), and a repeat head CT at 12 (+/-6) hours. ICH volumes were determined by blinded neuroradiologist reading, and a significant HE from baseline was considered ≥ 3 ml. Subjects were scanned with the device every 10 minutes for up to 72 hours.

Data from one subject was lost from operator error. Among the remaining nine, two experienced HE of ≥ 3ml (3 and 8.2 ml respectively). SENSE device readings were 100% concordant with the CT scan results.

## Introduction

Intracerebral hemorrhage (ICH) is a devastating form of stroke with mortality of 35 to 52%. Only 20% of survivors experience full recovery by 6 months (1), and ICH prevalence is expected to rise with the aging population. Current therapy is predominantly supportive (2), although there have been encouraging early results from neuro-interventional trials (3, 4) of stereotactic thrombolysis and acute blood pressure control (Shi 2017). Other approaches include hemostatic agents and coagulation factors with mixed results to date (5).

Hemorrhage expansion (HE) in ICH is associated with worse neurologic outcome and increased mortality (6). Davis et al found that for every 1 ml increase in ICH volume, patients were 7% more likely to worsen from independence towards requiring assistance, and from assistance to poor outcome. HE is not uncommon, occurring in about 38% of ICH patients within 20 hours (7).

Currently, HE is detected by repeat neuroimaging with CT or MRI, and can lead to significant changes in patient management such as surgical intervention (8).

Monitoring for HE after ICH is currently performed by clinical neurological exam and periodic neuroimaging. Clinical exam can be problematic in patients that are sedated and/or intubated. The exam can also lag serious intracranial events. In addition, at most institutions, the patient must be transported to radiology for the imaging which can take substantial time and risk. Finally, none of these techniques can be considered a continuous monitor of the brain and the hemorrhage. There are no current techniques or devices to provide continuous, non-invasive, bedside ICH monitoring in clinical practice.

The SENSE (Sensor Evaluation of Neurological Status in Emergencies) device was developed to address this clinical need. SENSE is a radiofrequency (RF) based technique utilizing the differential scattering and absorption of radio waves from intracranial tissues of varying dielectric constant e and conductivity a (9). Specifically, blood differs substantially in its electrical properties as compared with the grey and white matter of the brain (10). An RF signal incident on a blood collection of differing and from the surrounding tissue induces a time oscillating electric dipole moment in the blood collection (11). This results in a signal generated by the blood collection at the same frequency f of the incident signal, but differing in phase and amplitude, depending on the differing electrical characteristics, location of the blood, and its volume. These combined signals can be measured, and used to determine the presence or absence of significant changes in blood volume over time.

The use of microwaves as a potential medical imaging modality has been under development for some time (Semenov 2009). In microwave tomography (MT), microwave signals are scattered from biological tissues. The scattered signal amplitudes are used as input data to invert Maxwell’s equations, and obtain as a function of position within the tissue. Limitations include low dielectric contrast between some soft tissues, and mathematical complexity in solving these equations to obtain. Other approaches such as AI based analysis have been used in interpreting the scattered signals (Persson 2014) in more focused applications, such as the detection of acute stroke. Overall, these techniques have been focused on imaging or acute diagnosis of general brain pathology, and not as a monitor of changes in such pathology.

Korfhagen et al (Korfhagen 2013) demonstrated that a 400 MHz RF signal could detect as little as 1 ml of blood in a validated *in-vitro* brain gel model. They also found that the RF signal scattering was dependent on the geometry of the experimental blood collection. Kandadai et al (Kandadai 2014) demonstrated RF detection of 3 ml of blood in a porcine ICH preparation over a frequency range of 0.4-1 GHz. In addition, they found that the optimal RF frequency for intracranial blood detection was within 0.75 – 1 GHz range.

The overall goal of this study was to determine if SENSE could be used safely to monitor and detect ICH expansion in the Neurocritical Care unit. The secondary objective was to determine if SENSE could detect hemorrhage expansion of 3 ml or greater in these patients.

## Materials and Methods

### Study Objectives

The overall objective of this prospective, observational, single site, first-in-human pilot study of the SENSE device was to determine if SENSE could safely monitor ICH in the clinical environment. Up to 10 patients could be enrolled. Secondary objectives included human factors information to optimize the design, and operator challenges in the clinical environment. The results of the study were intended to guide further design changes for a planned pivotal clinical trial of the device.

The primary endpoint for this early feasibility study was the correlation of SENSE with CT changes over time. An expansion of 3 ml or more in a given ICH was considered significant.

### Study Population

The study population was patients presenting with a diagnosis of spontaneous ICH to a single urban comprehensive stroke center (CSC). Emergency Department based Clinical Research Coordinators (CRC) screened for ICH patients at the study site. Patients were enrolled within 24 hours of symptom onset.

In order to be a candidate for the study, ICH patients were 22 years of age or older, with a diagnostic head CT performed within 24 hours of symptom onset, an ICH volume greater than or equal to 1 and less than 90 ml, and able to participate in study procedures. Potential subjects were excluded if pregnant or lactating, had a history of seizure disorder or were on medications that lowered the seizure threshold, had an intraventricular hemorrhage (IVH), subdural hemorrhage (SDH), epidural hemorrhage (EDH), or secondary cause of the ICH, if surgical ICH evacuation or continuous EEG monitoring was planned within 24 hours, or if taking medications that increased hemorrhage risk.

Patients were approached by CRCs after the screening process, and enrolled after appropriate written informed consent was obtained from the patient, or approved legal surrogate. Following subject enrollment, the SENSE device was placed on the patient for up to 72 hours. The device performed a scan every 10 minutes, and an individual scan took about 40 seconds. The SENSE data was recorded internally in the device, and later downloaded by study personnel after the monitoring period was complete for a given subject. The treating clinicians were blinded to the SENSE data.

For each subject, data recorded included basic subject demographic data, past medical history, and radiologist’s interpretation of the neuroimaging studies. Adverse events (AE) and serious adverse events (SAEs) were noted and recorded.

The study protocol was approved by the FDA under and IDE and by the local IRB.

### Imaging Protocol

The following CT imaging protocol was followed for each subject:

- Diagnosis: CT Scan 1 – standard of care; diagnostic head CT scan within 24 hours of symptom onset.
- First Study CT: CT Scan 2 – study CT; first study CT scan as soon as practicable after informed consent, followed by SENSE monitoring within 15 minutes of completion of the first CT scan.
- 12 Hours: CT Scan 3 – standard of care; CT scan 12 (±6) hours after first study CT scan.
- 72 Hours: CT Scan 4 – study CT; end of study CT scan 72 (±12) hours after first study CT scan.

Study eligibility ICH volumes were calculated from the diagnostic CT by clinical and/or research staff using the ABC method (12). Subsequently, ICH and edema volumes were determined by a neuroradiologist blinded to the clinical and SENSE device data. Quantification of volumes were performed using computer-assisted methods. The blinded evaluator was required to place seed-points within the volume of interest and adjust lower and upper intensity Hounsfield unit (HU) thresholds until the entire volume was correctly selected.

### SENSE Device

The SENSE device was comprised of two segments; a headset worn by the patient (Figure 1), and the controlling electronics (not shown). These two items were connected by a semi-rigid cable. The headpiece contained 9 circular microwave patch antennae with an operating frequency of 913 MHz. The total power radiated for a given antenna when active was 1 mW.

**Figure 1:**
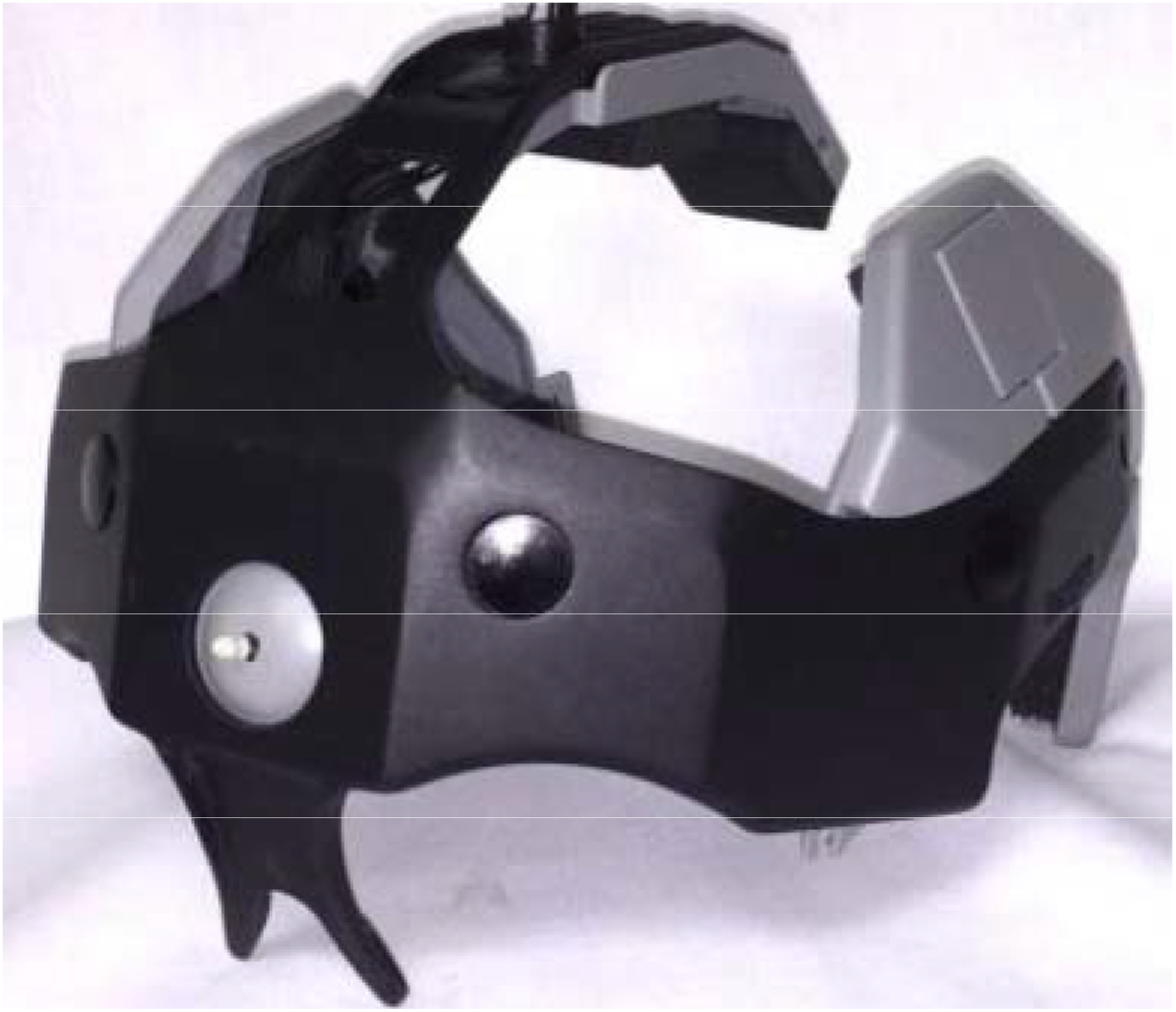
SENSE headset piece (controlling electronics not shown). The antenna array (9 total) is contained within the headpiece, along with appropriate wiring. The antenna signals are input and output via a cable (not shown) that communicates with the control box.

The antenna array was configured as 3 sagittal arrays of 3 antenna each; one in the midline of the patient’s head, and the remaining on the left and right sides respectively. The antennae were offset from direct contact with a given subject’s head by a foam insulating pad. All antennae were approximately oriented radially towards the center of the intracranial region. All patient-contacting materials and electronics underwent and passed approved biological and electronic safety testing prior to use.

For a given measurement, one antenna would transmit the incident signal, and the remaining 8 received the altered signal as modified by absorption and scattering from intracranial tissues. In a single measurement, all 9-antenna served as the transmitting antenna, thus yielding 72 total measurements. For a cycle, the measurement would be repeated for 10 iterations. A single cycle took about 40 seconds. The data were stored on a secured internal hard drive of the driving electronics, and were downloaded by study personnel once data acquisition for the given patient was completed. Patients were scanned every 10 minutes during the study period.

### SENSE Data

Methods used in the SENSE algorithm were developed from in vitro and in vivo porcine experiments (9, 13). First, for each pair of antennae, the raw voltage data was compared to the first, or baseline, measurement. This value was denoted ΔTR, where T is the transmitting antenna and R is the receiving antenna. Then, for this time point, delta values for all pairs were compared. By comparing the delta values to a set of threshold values, the change was determined to be due to a change in the ICH volume, edema development, movement, or system drift. While the SENSE device can detect changes due to both increases and decreases (due to clot retraction) in hemorrhage size, only increases in hemorrhage volume greater than 3 mL were considered significant and were denoted as active bleeding.

## Results

### Subjects

A total of 10 patients were enrolled from May through October 2017, and 9 completed the protocol. The average age was 57 years (range: 43-70), and there were 5 female subjects. Details of the ICH locations and volumes are shown in Table 1. Patients were monitored with SENSE for an average of 60 (range: 23-72) hours. There were no Serious Adverse Events. Adverse events were predominantly minor skin and nose irritation from the nosepiece of the headset.

**Table 1:** ICH Characteristics

| ICH Location | Initial V (ml) | V (12 h) | V (72 h) | 12 h ICH Expansion |
| --- | --- | --- | --- | --- |
| R BG,Thalamus | 1 | 1 |  |  |
| L BG, Thalamus, IC | 3.8 | 3.8 | 3.2 |  |
| L Lobar | 20.2 | 20.6 | 18 |  |
| BG | 22 | 25 | 20 | Y |
| R Lobar | 35.1 | 33.4 | 30.4 |  |
| L BG, thalamus, IC | 24.8 | 23.5 | 18.3 |  |
| R BS, Cerebellar | 12.7 | 20.9 |  | Y |
| L BG | 12.3 | 11.3 | 11.2 |  |
| L Lobar | 61.4 | 59.5 | 51.7 |  |
| L BG, Thalamus, IC, Caudate Nucleus | 51.5 | 50.4 | 48.9 |  |

### CTs

The initial head CT for these patients showed an average ICH volume of 24.5 (SD 19.7) ml. Of the 10 patients, 9 completed the protocol. The SENSE data for one patient was lost due to operator error of the device. Of the remaining 9 patients, 2 had a significant expansion of 3.0 and 8.2 ml, respectively. The expansion for both patients occurred within 12 hours of onset, and was demonstrated on the 12-hour study CT.

### SENSE

As shown in Table 1, the SENSE data was 100% concordant with the CT results. The SENSE signals showed significant changes in time for the two expansions, and did not show any such changes for the 7 patients who did not experience a hemorrhage expansion.

## Discussion

In summary, the SENSE device was used to safely monitor ICH patients in a clinical environment. In addition, it was able to detect HE of ≥ 3 ml in this prospective, single-site pilot trial. Significant limitations include the small sample size n of 10 patients, and the single-site limits the generalizability of our results. However, these results are promising, and certainly warrant further study.

Currently, ICH patients are monitored by serial neurologic exam, and neuroimaging. In addition, intracranial pressure monitoring (ICP) is sometimes utilized in a subset of patients (Tuan 2013). However to the authors’ knowledge, there are no non-invasive monitors of ICH in current clinical use. In addition, the clinical exam may be difficult in a sedated and/or paralyzed and intubated patient such that a neurological exam is not possible. Also HE can occur in such patients, and may not be detected for hours, depending on the schedule of neuroimaging. There is clearly a clinical need for such a monitor, and the results presented here suggest that RF monitoring may be a useful approach.

## Data Availability

Available data are presented in the manuscript

## Author Contributions Statement

All authors planned and designed the study. JJ wrote the grant, obtained the data, and performed the data analysis. GJ performed data analysis and drafted the manuscript. OA and ML assisted in grant preparation, and reviewed the manuscript. WK assisted in study design, served as site PI, and reviewed the manuscript.

